# “Mortality Prediction in Heart Failure using Explainable AI: Development and Validation of the EXACT-HF Model in a Nationwide Multi-Ethnic Asian Cohort”

**DOI:** 10.64898/2026.09.10.26362800

**Authors:** Silingga Metta Jauhari, Wesley Yeung, Tony Li, Joey Chua, Audrey Zhang, Lim Eng How, Robin Cherian, David Sim, Sheldon Lee, Dinna Soon, Chan Po Fun, Loh Seet Yong, Fazlur Jaufeerally, Siew Pang Chan, Mark Chan, James Yip, Lin Weiqin, Raymond Wong

## Abstract

**Background:** Established heart-failure (HF) risk scores were developed largely in Western cohorts, assume linear predictor effects, and stop at prognosis without indicating action. We developed and externally validated machine-learning (ML) models for 30-day and 1-year mortality in a multi-ethnic Asian HF population, benchmarked against three established scores, and tested whether they localised correctable guideline-directed medical therapy (GDMT) gaps.

**Methods:** We analysed 9,050 adults hospitalised with HF across eight Singapore hospitals (November 2016–March 2023). Gradient-boosted trees (XGBoost, primary) and elastic-net logistic regression were developed on three hospitals (n=4,034) and validated on five independent hospitals (n=5,016), without patient or site overlap. Comparators (MAGGIC, Singapore HF Score, OPTIMIZE-HF-Asia) were logistically recalibrated on development data.

**Results:** On external validation XGBoost achieved AUROCs of 0.862 (95% CI 0.833–0.890) for 30-day and 0.749 (0.734–0.764) for 1-year mortality, versus 0.711–0.754 and 0.682–0.699 for the three recalibrated scores; logistic regression performed near-identically (0.864, 0.757) and was better calibrated at 1 year. Superiority held under temporal validation, multiple imputation and optimism correction, although the 30-day advantage was no longer distinguishable from MAGGIC once medications were removed. Among predicted-high-risk patients with reduced ejection fraction, mean GDMT exposure was 0.92 of four pillars versus 1.87 across all HFrEF patients, and 36.3% received none of the four versus 2.0% of low-risk patients. Ten variables reproduced full-model 30-day discrimination (AUROC 0.865).

**Conclusions:** In external validation, interpretable models substantially outperformed established HF risk scores for short- and long-term mortality and simultaneously localised correctable therapeutic gaps, supporting a prediction-to-action tool deployable with ten bedside variables.

**Clinical Perspective:** *What Is New?:*

- In 9,050 patients hospitalised for heart failure across eight Singapore hospitals, prediction models estimated 30-day and 1-year all-cause mortality substantially better than three established risk scores (30-day AUROC 0.86 versus 0.71–0.75), validated in five hospitals independent of model development.
- Ten routinely recorded bedside variables reproduced the full 85-variable model’s 30-day discrimination. This enables bedside risk estimation without electronic health record integration.

*What Are the Clinical Implications?:*

- The same model identifies both who is at risk and which guideline-directed therapies they lack: flagged patients were receiving a mean of 0.9 of the four pillars, against 1.9 across all patients with reduced ejection fraction, although most non-prescription had a documented renal or haemodynamic contraindication, so this correctable fraction is an upper bound requiring prospective validation.

## Introduction

Heart failure (HF) remains among the most lethal and resource-intensive chronic cardiovascular syndromes^1^, with in-hospital and early post-discharge mortality that rivals many malignancies. HF outcomes also vary across ethnic groups within Asia^2^, reflecting known health inequalities in HF mortality and guideline-directed therapy uptake by ethnicity and sex; prediction tools trained on non-representative cohorts risk perpetuating rather than correcting these disparities, underscoring the need for risk-prediction tools developed and validated in multi-ethnic Asian populations rather than extrapolated from Western cohorts. Accurate, individualised mortality estimation is central to shared decision-making, triage of intensified surveillance, timing of advanced therapies, and enrolment into transitional-care programmes^3^. It is equally central to a health-system question that pure prognostication rarely answers: among the patients a model flags as high risk, how many carry a modifiable, guideline-addressable cause of that risk.

Several risk scores are in routine use. The Meta-Analysis Global Group in Chronic Heart Failure (MAGGIC) score is the most widely validated^4^, while region-specific instruments such as the Singapore HF Score^5^ and OPTIMIZE-HF-Asia^6^ were derived to better reflect Asian populations^7^. These tools share three structural limitations^8^. First, they were developed predominantly in cohorts that predate the contemporary four-pillar era of guideline-directed medical therapy (GDMT), in which angiotensin receptor–neprilysin inhibitors (ARNI) and sodium–glucose co-transporter-2 inhibitors (SGLT2i) have materially altered outcomes. Second, they assume that each prognostic variable contributes linearly and additively, an assumption that is biologically implausible for markers such as serum sodium and urea, where risk rises steeply beyond physiological thresholds. Third, they output a probability but no action: a high MAGGIC score identifies a patient likely to die, not a lever a clinician can pull.

Machine-learning (ML) methods can capture nonlinear and interaction effects without pre-specification, and modern interpretability techniques (e.g. SHapley Additive exPlanations, SHAP) make their predictions inspectable rather than opaque^9^. Prior ML HF-mortality models have, however, frequently been limited by single-centre design, internal-only validation, comparison against poorly calibrated benchmarks, or the absence of any actionability layer^9–13^, leaving clinicians a more accurate but equally inert risk number.

We therefore developed and externally validated ML models for 30-day and 1-year all-cause mortality in a large, multi-ethnic Asian HF cohort spanning eight hospitals. Our objectives were to (i) benchmark ML discrimination and calibration against three established, fairly recalibrated risk scores in a fully independent five-hospital external cohort; (ii) subject any observed advantage to a comprehensive battery of robustness analyses (temporal validation, multiple imputation, optimism-corrected bootstrapping, a medication-free sensitivity model, and subgroup fairness assessment); and (iii) test whether the model can move beyond prognosis by quantifying correctable GDMT gaps among predicted-high-risk patients. The study is reported in accordance with TRIPOD+AI.

## Methods

### Study design, setting, and participants

We conducted a retrospective, multi-site cohort study using a harmonised Heart Failure Registry collected from electronic health records (EHR) covering eight Singapore hospitals over the period November 2016 to March 2023. Eligible participants were adults (age ≥18 years) hospitalised with a primary diagnosis of heart failure. Where a patient had multiple admissions, the most recent admission was retained (deduplication by national identifier), and analysis was performed at the level of the index baseline visit. Patients with a missing 30-day vital-status outcome were excluded. The final analytic cohort comprised 9,050 patients. Sample size was determined by the full available registry over the study period rather than by a priori power calculation, consistent with recommended practice for model development using large real-world registry data; the resulting development (n=4,034) and external validation (n=5,016) cohorts yielded 142 and 235 thirty-day deaths respectively, sufficient for the bootstrap-based confidence intervals reported throughout.

### Development versus validation separation

To emulate real-world transportability rather than a within-sample split alone, sites were partitioned a priori into a development set of three hospitals (NUH, NHC, CGH; n=4,034) and an external validation set of five independent hospitals (KTPH, NTFGH, SGH, TTSH, WHC; n=5,016). There was no patient or site overlap between the two. Within the development set, an 80/20 stratified train/test split (stratified on 30-day mortality; n=3,227 train, n=807 test) supported model tuning and internal evaluation.

### Outcomes

The two pre-specified outcomes were all-cause mortality at 30 days and at 365 days from the index admission, ascertained from registry death records (death date versus admission date) with administrative censoring at the study end. Vital status is not subject to assessor blinding.

### Candidate predictors

Predictors were restricted to information available at or before discharge from the index admission, comprising demographics (age, sex, ethnicity, marital status, body-mass index, smoking, alcohol use), admission vital signs, presenting symptoms and NYHA class, documented comorbidities, relevant past medical history, admission laboratory values (full blood count, sodium, potassium, creatinine, urea, random glucose, eGFR), electrocardiographic QRS duration and left-ventricular ejection fraction, and discharge medications spanning the GDMT classes and adjuncts. The full (’all-variable’) model used 85 encoded features. Because discharge medications carry a well-recognised risk of confounding by indication, the eleven medication variables were held in a separate feature block so that they could be removed en bloc for a pre-specified sensitivity model.

### Missing data and preprocessing

Preprocessing statistics were fitted on the training data only and applied unchanged to test and external data. Continuous predictors were median-imputed and winsorised at the 0.1st–99.9th percentiles; categorical predictors received an explicit ‘Missing’ category and were one-hot encoded. Variable-level missingness is reported in Supplementary Table S1. The robustness of this strategy was tested against multiple imputation by chained equations (MICE, m=5) in sensitivity analysis.

### Model development

Two model families were developed. The primary model was a gradient-boosted decision-tree ensemble (XGBoost), with hyperparameters tuned by Bayesian optimisation (Optuna, TPE sampler, 30 trials, 5-fold cross-validated AUROC objective; search over number of estimators 200–800, learning rate 0.01–0.3, maximum depth 3–8, subsample and column-subsample 0.6–1.0, L2 regularisation 0.1–5.0). The final selected hyperparameters were 257 estimators, learning rate 0.0125, maximum depth 7, subsample 0.65, column-subsample 0.81, and L2 regularisation 3.76 for the 30-day model, and 784 estimators, learning rate 0.0111, maximum depth 8, subsample 0.73, column-subsample 0.77, and L2 regularisation 3.28 for the 1-year model (random_state=42, objective=binary:logistic throughout). The co-reported model was an elastic-net penalised logistic regression (LogisticRegressionCV, saga solver, L1 ratio 0.5, 5-fold cross-validation on standardised features). Gradient boosting was selected as the primary model because it natively captures the nonlinear threshold effects and interactions that motivate moving beyond linear scores, and because it supports patient-level SHAP interpretability; logistic regression is reported alongside as a transparent, easily deployed reference. No class-reweighting or resampling was applied to correct for the low ( ∼4%) 30-day event rate; imbalance was instead addressed by prioritising calibration-sensitive metrics (Brier score, calibration slope and intercept) alongside discrimination throughout evaluation, since resampling-based correction can itself distort calibration.

### Comparator risk scores

Three established scores were computed: MAGGIC, the Singapore HF Score, and OPTIMIZE-HF-Asia. Raw scores are not calibrated probabilities; naïve min–max rescaling produced grossly miscalibrated risks (e.g. MAGGIC Brier ≈0.31). To provide a fair comparator, each score was recalibrated by fitting a single-variable logistic recalibration on its underlying linear predictor using the development data only. Recalibrating on the linear predictor (rather than the already-sigmoid-squashed probability) was essential for numerical stability, particularly for the heavily right-skewed OPTIMIZE-HF-Asia output.

### Evaluation

Discrimination (AUROC, AUPRC), calibration (Brier score, calibration slope and intercept), and threshold-based operating characteristics (at the Youden point) were evaluated on the internal test set and, primarily, on the pooled external cohort and each of the five external hospitals. Uncertainty was quantified by outcome-stratified bootstrap (2,000 resamples) 95% confidence intervals. Clinical usefulness was assessed by decision-curve analysis (net benefit across threshold probabilities) and by reclassification versus recalibrated MAGGIC (continuous and categorical net reclassification improvement [NRI] and integrated discrimination improvement [IDI]). Kaplan–Meier analysis across model-defined risk tertiles assessed survival separation.

### Robustness, fairness, interpretability, and actionability

Pre-specified robustness analyses comprised: (i) temporal validation, training on pre-2021 admissions and testing on 2021-onward admissions to probe stability across the ARNI/SGLT2i uptake era; (ii) multiple-imputation sensitivity; (iii) optimism-corrected internal performance by bootstrap, evaluated on out-of-bag rows (the textbook full-sample evaluation was rejected because tree ensembles can near-perfectly memorise resampled training rows, spuriously understating optimism); and (iv) the medication-free (’no-treatment’) model. Fairness was assessed by subgroup discrimination, calibration, and error rates across age, sex, ethnicity, and ischaemic aetiology, with event counts reported so that unstable small-subgroup estimates are visible. Interpretability used SHAP importance and dependence plots. Actionability was quantified by measuring, among predicted-high-risk patients with reduced ejection fraction (LVEF <40%), the proportion missing each of the four GDMT pillars. A minimal-feature model was built by adding SHAP-ranked predictors sequentially to identify the smallest bedside variable set matching full-model performance.

Analyses used Python^14^ (scikit-learn^15^, XGBoost^16^, Optuna, SHAP). The study was conducted under the governing registry ethics approval; reporting follows TRIPOD+AI.

## Results

### Cohort characteristics

The analytic cohort of 9,050 patients (4,034 development, 5,016 external validation) had a mean age of 70.2 years and was 70.3% male; 64.3% were Chinese, 21.7% Malay, and 11.5% Indian2. Reduced ejection fraction (LVEF <40%) was present in 71.9%, with a mean LVEF of 27.6%. Thirty-day mortality was 4.2% overall (development 3.5%, external 4.7%) and 1-year mortality 22.6% (development 20.7%, external 24.1%). Development and external cohorts were broadly similar clinically, though the external hospitals had modestly older patients, higher creatinine, and lower uptake of the newer GDMT agents (ARNI 8.2% vs 23.4%; SGLT2i 8.0% vs 16.6%), reflecting real inter-hospital practice variation and reinforcing the value of geographically external validation (Table 1; Figure 1). Because discharge medications were among the strongest predictors, this prescribing difference means the external cohort tested transportability under a materially different treatment distribution from the one the model was trained on.

**Figure 1.**
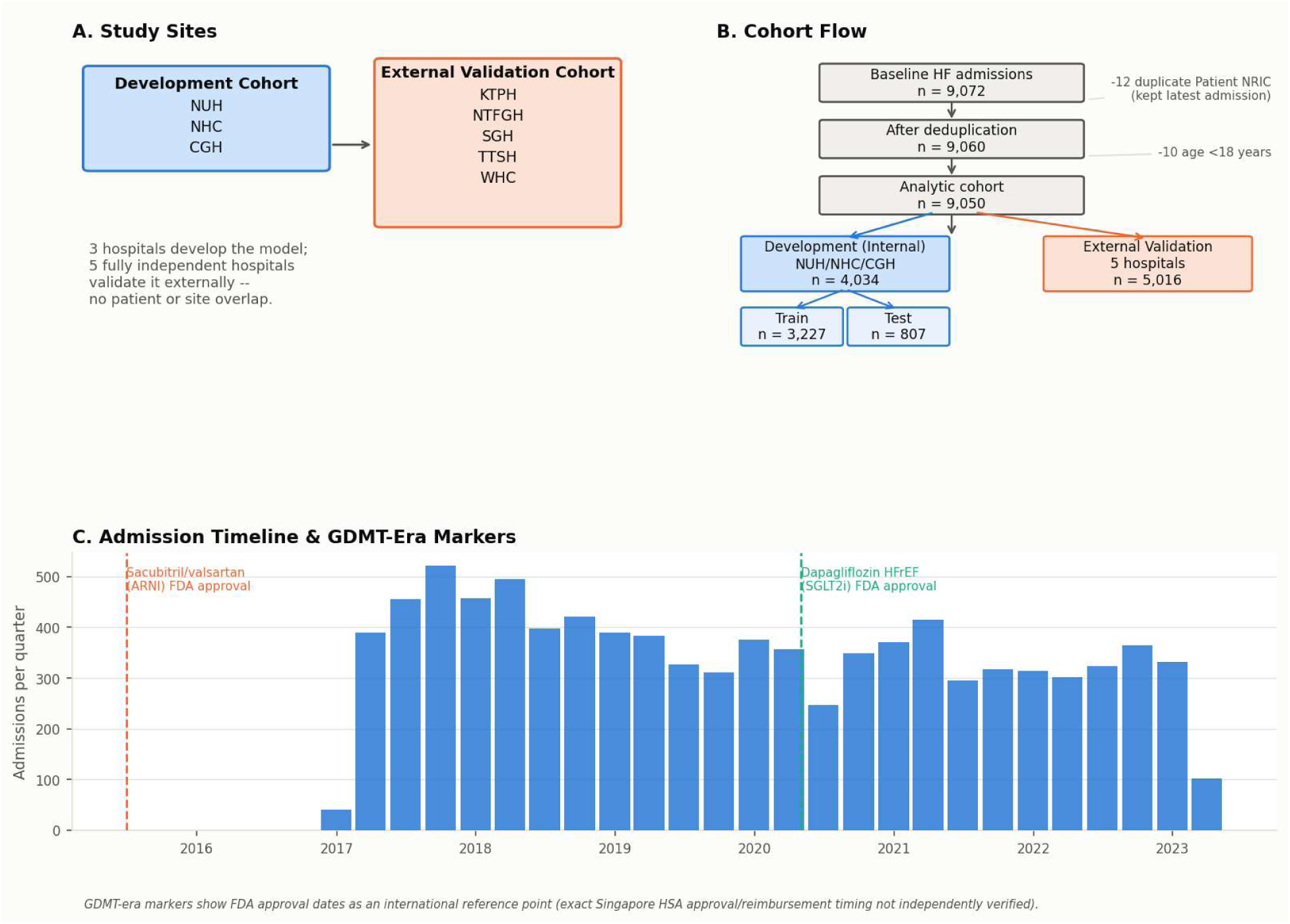
Study Design, Cohort Flow, and Contemporary GDMT Era. Cohort design and study flow. Eight hospitals were partitioned a priori into a three-hospital development set (internal 80/20 train/test split) and a fully independent five-hospital external validation set, with no patient or site overlap.

**Table 1.**
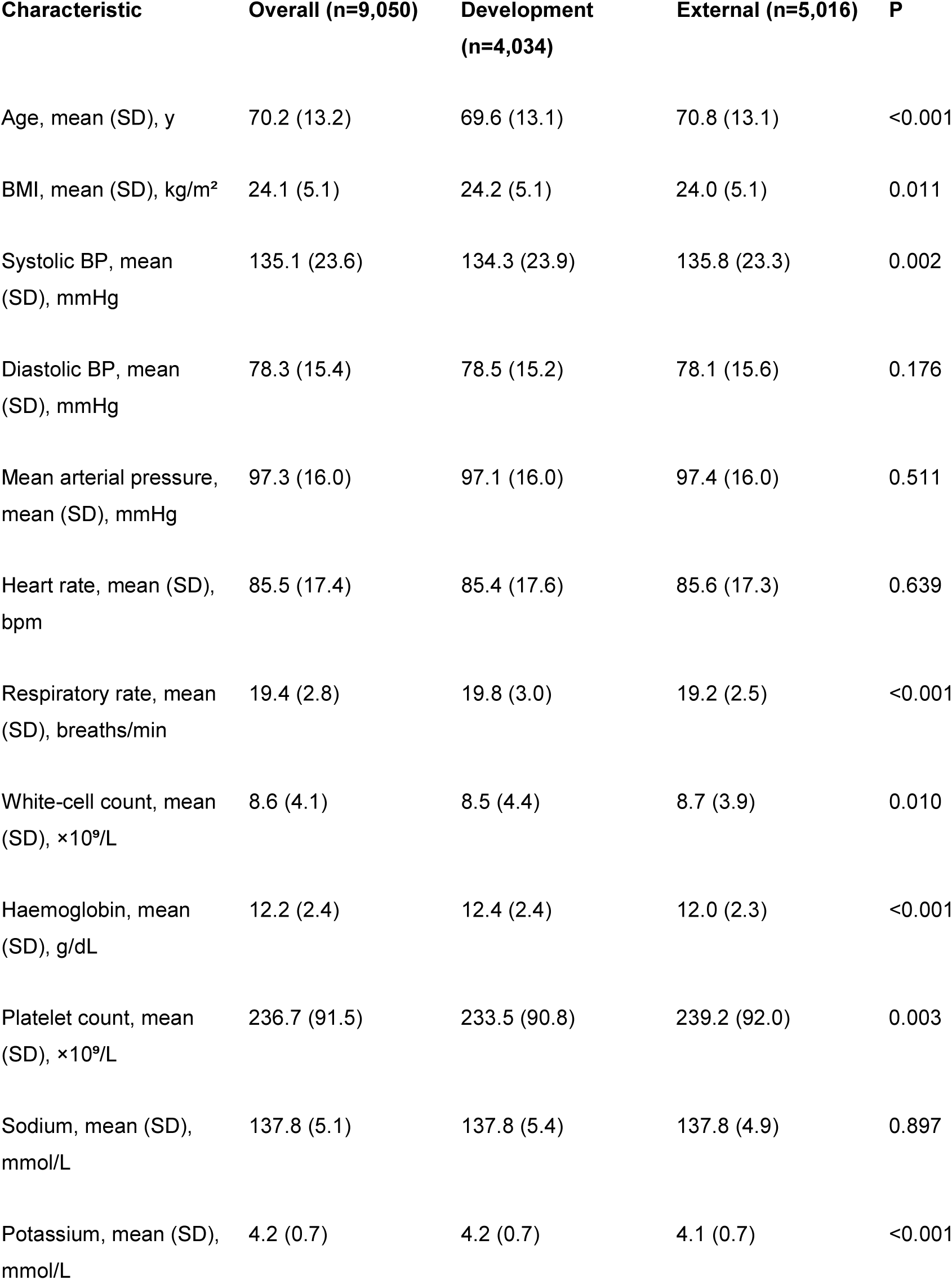
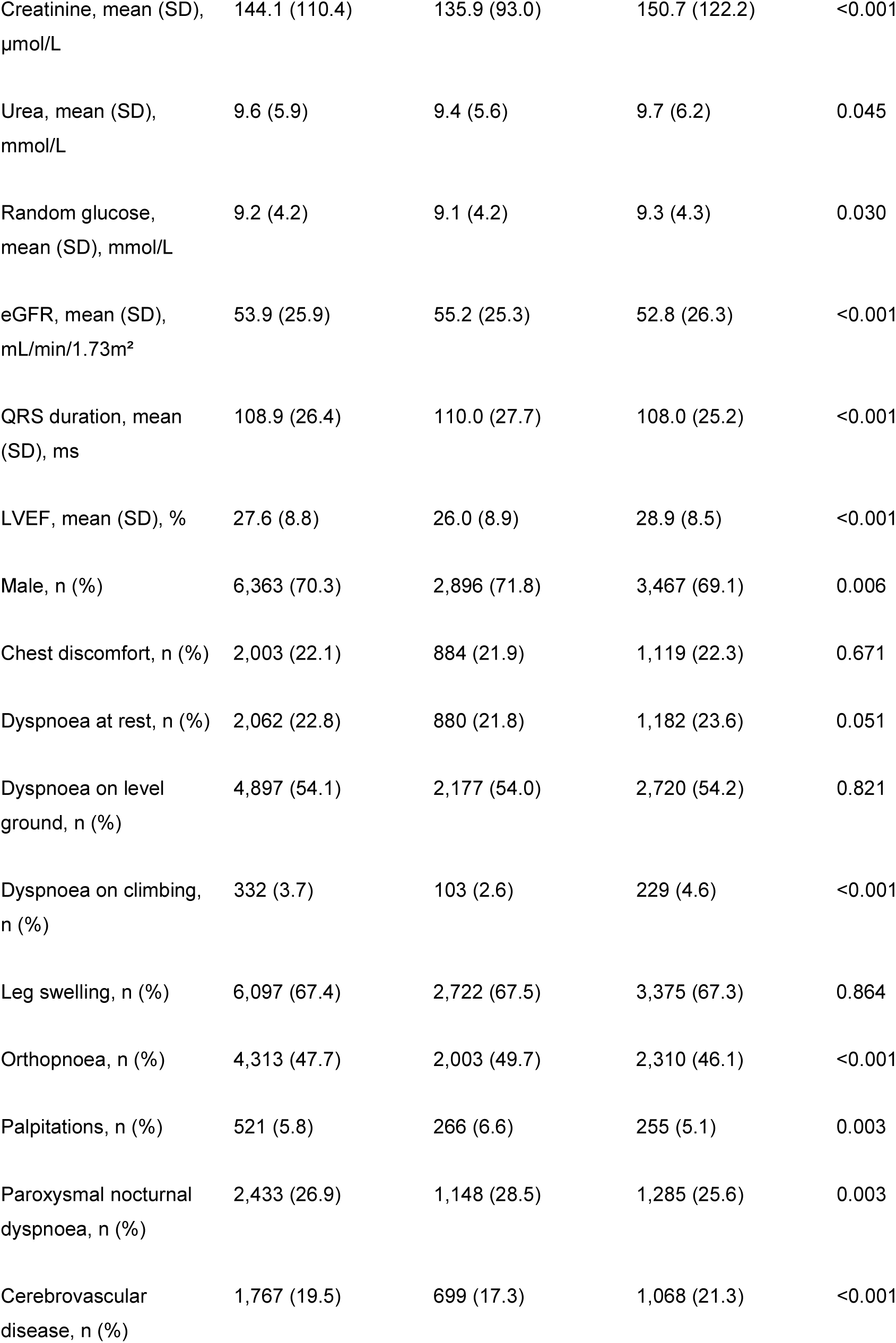
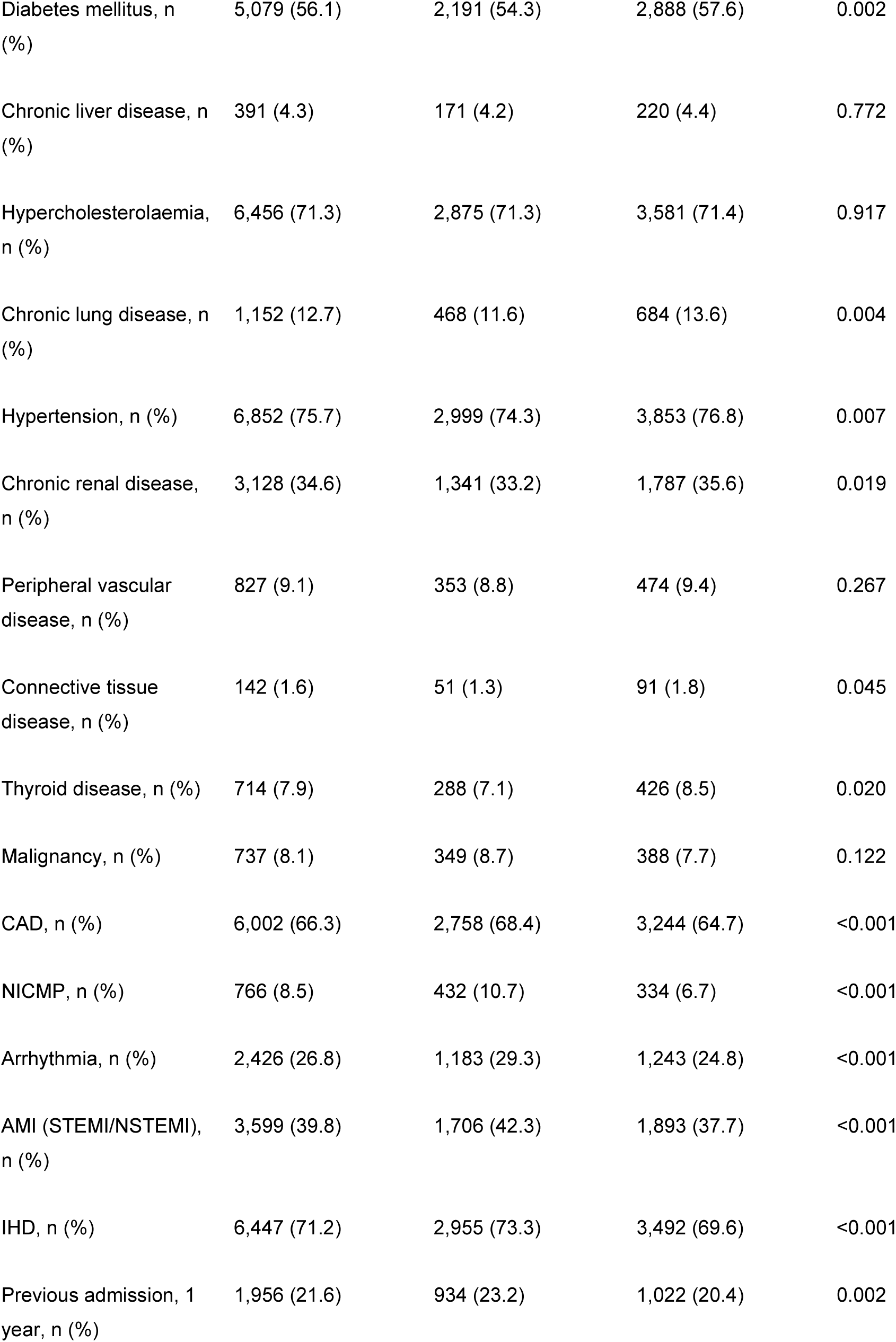
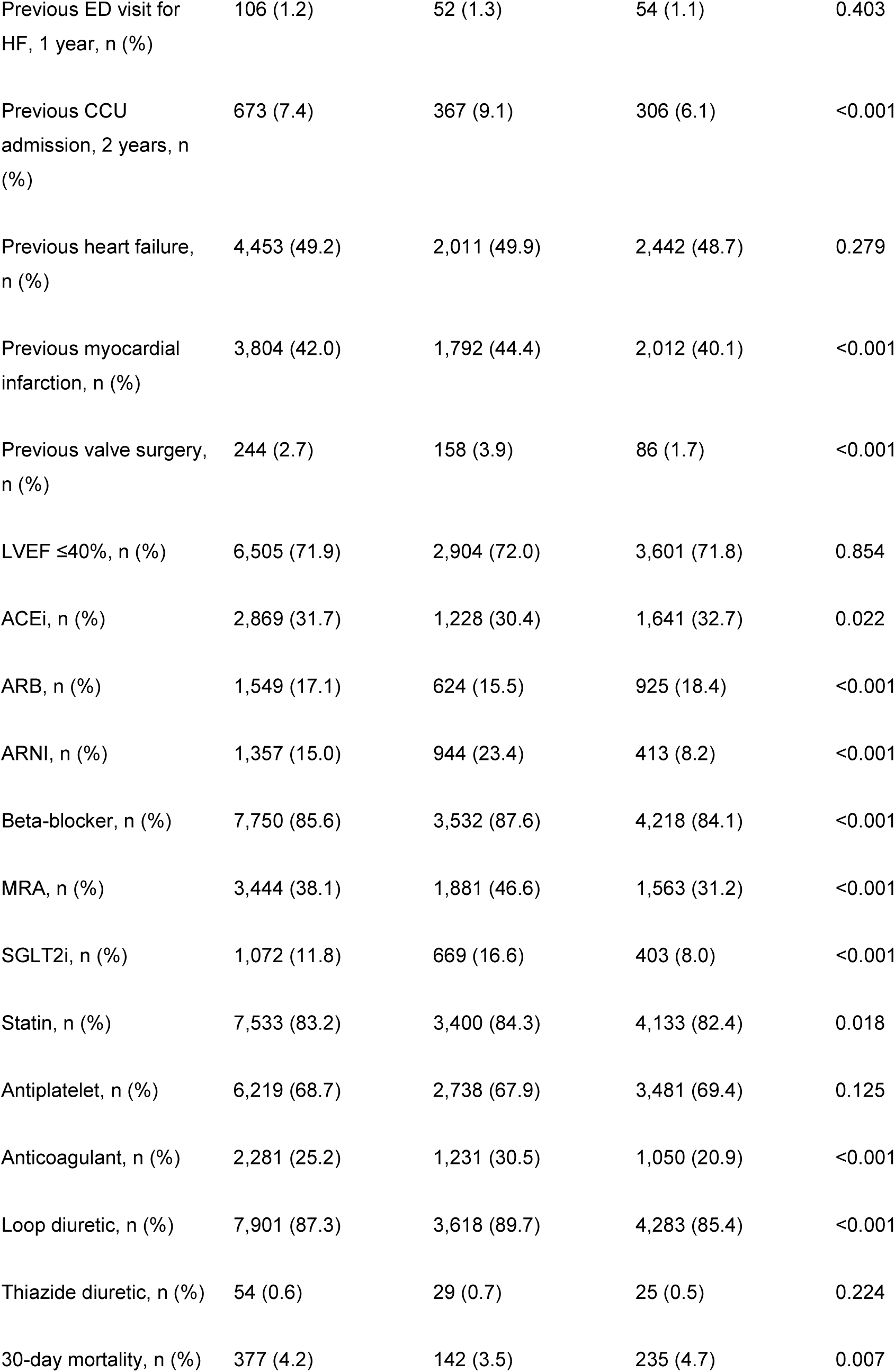
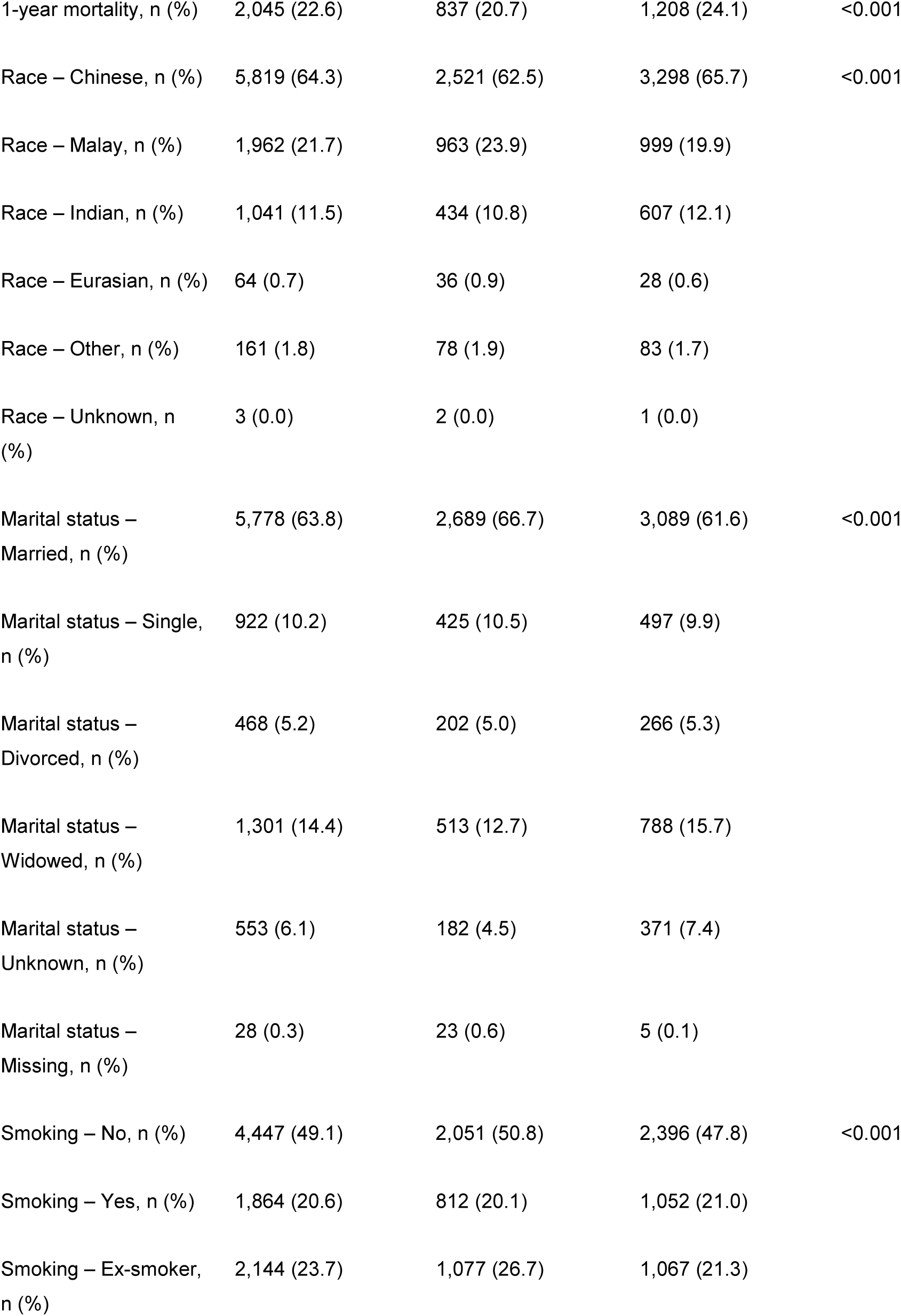
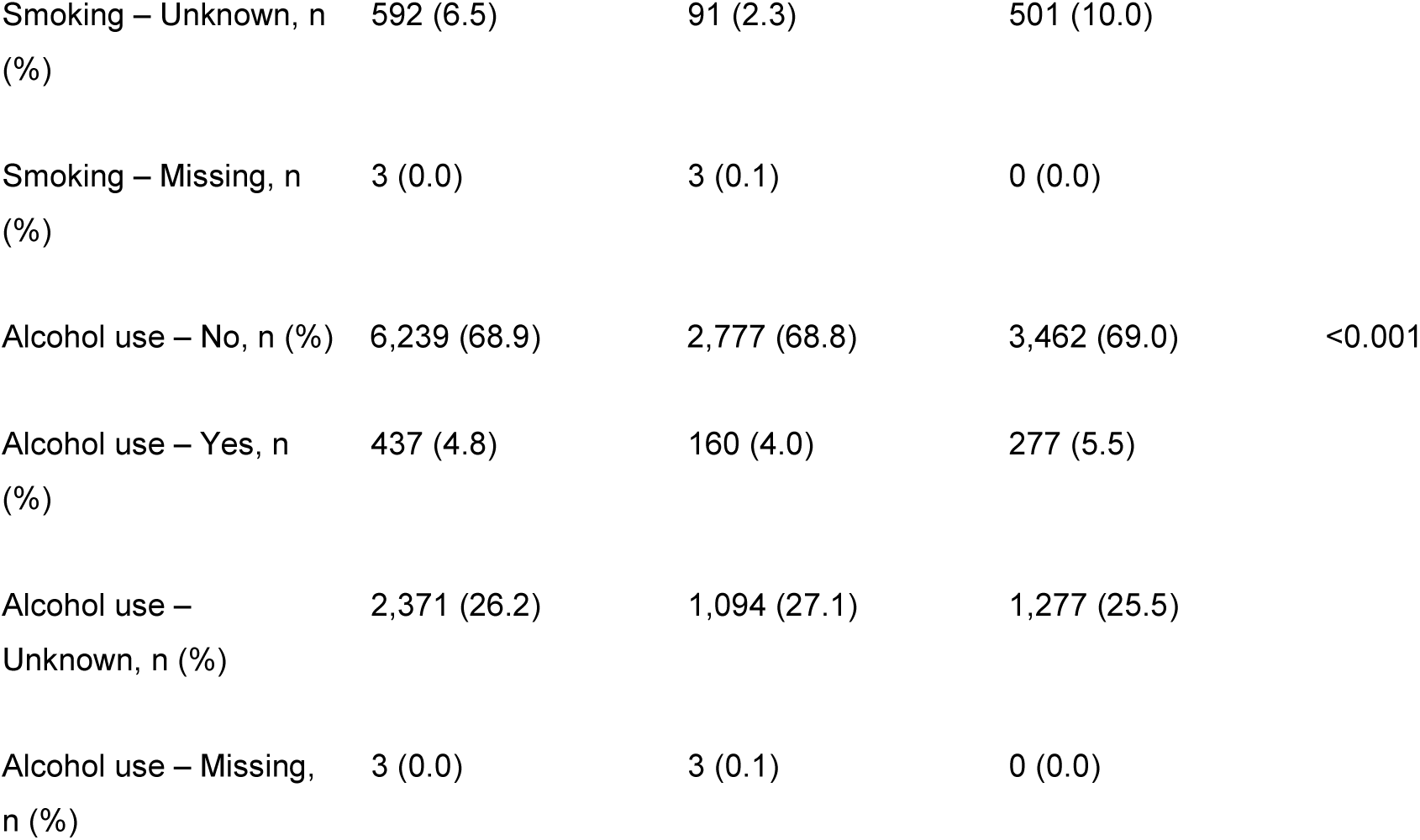
Baseline characteristics of the development and external validation cohorts. **Per-variable missingness prior to multiple imputation is provided in Supplementary Table S1.** BP, blood pressure; BMI, body-mass index; LVEF, left-ventricular ejection fraction; MRA, mineralocorticoid-receptor antagonist; ARNI, angiotensin receptor–neprilysin inhibitor; SGLT2i, sodium–glucose co-transporter-2 inhibitor. Baseline characteristics were tabulated using the tableone Python package^25^. P values compare the development and external cohorts and are reported for completeness only; at this sample size they detect clinically trivial differences, and standardised mean differences are the more informative measure of cohort comparability.

### Discrimination and calibration on external validation

On the pooled external cohort, the primary XGBoost model discriminated 30-day mortality with an AUROC of 0.862 (95% CI 0.833–0.890) and 1-year mortality with an AUROC of 0.749 (0.734–0.764). The co-reported logistic-regression model performed near-identically (0.864 and 0.757). Both ML models were well calibrated at 30 days (Brier 0.030–0.031), although calibration slopes modestly exceeded unity (XGBoost 1.172, 95% CI 1.076–1.279; logistic regression 1.112, 1.014–1.221). At 1 year Brier scores were comparable (0.152–0.160) but the two models diverged in calibration: logistic regression was well calibrated (slope 0.986, 95% CI 0.911–1.060) whereas XGBoost systematically over-dispersed predicted risk (slope 0.715, 0.662–0.770), a pattern consistent across all five external sites (site-specific slopes 0.63–0.79). Logistic regression is therefore the better-calibrated model at one year. By contrast, the three established scores, even after fair recalibration, discriminated substantially less well: 30-day AUROC 0.754 (MAGGIC), 0.732 (OPTIMIZE-HF-Asia), and 0.711 (Singapore HF Score); and 1-year AUROC 0.682–0.699 (Table 2; Figure 2). Discrimination was consistent across the five external hospitals individually (30-day AUROC range 0.855–0.901; 1-year 0.716–0.757), while calibration slopes varied more by site (30-day 0.92–1.22; 1-year 0.63–0.79); the smallest site (n=67, 6 thirty-day events) had correspondingly wide confidence intervals and should be interpreted cautiously (Supplementary Table S9). The ML advantage over the best comparator (MAGGIC) was approximately 0.11 AUROC at 30 days and 0.05 at 1 year.

**Figure 2.**
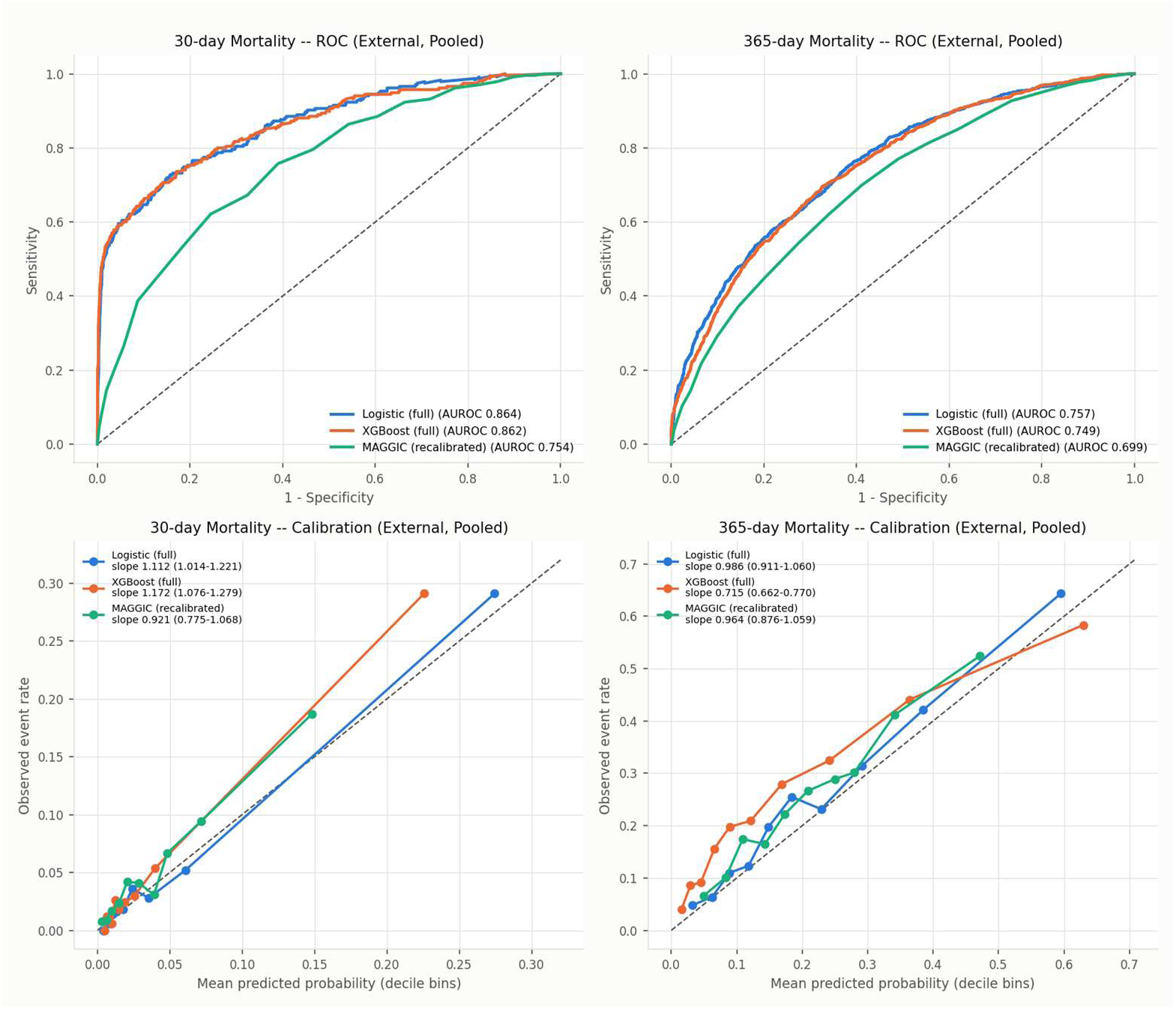
Discrimination and Calibration (External Validation. Discrimination (ROC curves) and calibration on external validation for XGBoost, logistic regression, and the three recalibrated comparator scores, for 30-day and 1-year mortality.

**Table 2.** Discrimination and calibration on external validation (five independent hospitals, n=5,016) Comparator scores were recalibrated on their linear predictor using development data only. 95% CIs are outcome-stratified bootstrap (2,000 resamples). Perfect calibration corresponds to slope 1 and intercept 0.

**30-day mortality (235 events)**
| Model | AUROC (95% CI) | Brier | Calibration slope (95% CI) |
| --- | --- | --- | --- |
| XGBoost (primary) | 0.862 (0.833–0.890) | 0.030 | 1.17 (1.08–1.28) |
| Logistic regression | 0.864 (0.836–0.891) | 0.031 | 1.11 (1.01–1.22) |
| MAGGIC (recalibrated) | 0.754 (0.721–0.785) | 0.042 | 0.92 (0.78–1.07) |
| OPTIMIZE-HF-Asia (recal.) | 0.732 (0.701–0.762) | 0.044 | 2.26 (1.88–2.64) |
| Singapore HF Score (recal.) | 0.711 (0.680–0.742) | 0.044 | 2.03 (1.71–2.37) |

| Model | AUROC (95% CI) | Brier | Calibration slope (95% CI) |
| --- | --- | --- | --- |
| XGBoost (primary) | 0.749 (0.734–0.764) | 0.160 | 0.72 (0.66–0.77) |
| Logistic regression | 0.757 (0.741–0.772) | 0.152 | 0.99 (0.91–1.06) |
| MAGGIC (recalibrated) | 0.699 (0.683–0.715) | 0.168 | 0.96 (0.88–1.06) |
| Singapore HF Score (recal.) | 0.685 (0.668–0.702) | 0.171 | 0.87 (0.76–0.98) |
| OPTIMIZE-HF-Asia (recal.) | 0.682 (0.664–0.699) | 0.170 | 0.89 (0.77–1.02) |

### Clinical utility and reclassification

Across clinically relevant threshold probabilities, both ML models delivered positive net benefit exceeding treat-all and treat-none strategies and exceeding recalibrated MAGGIC (Figure 3). Relative to recalibrated MAGGIC, XGBoost improved 30-day risk classification with a continuous NRI of 0.556 (95% CI 0.429– 0.678), a categorical NRI of 0.206 (0.136–0.275), and an IDI of 0.224 (0.189–0.259); corresponding 1-year figures were a continuous NRI of 0.534 (0.470–0.596) and IDI of 0.086 (0.075–0.097) (Table 3).

**Figure 3.**
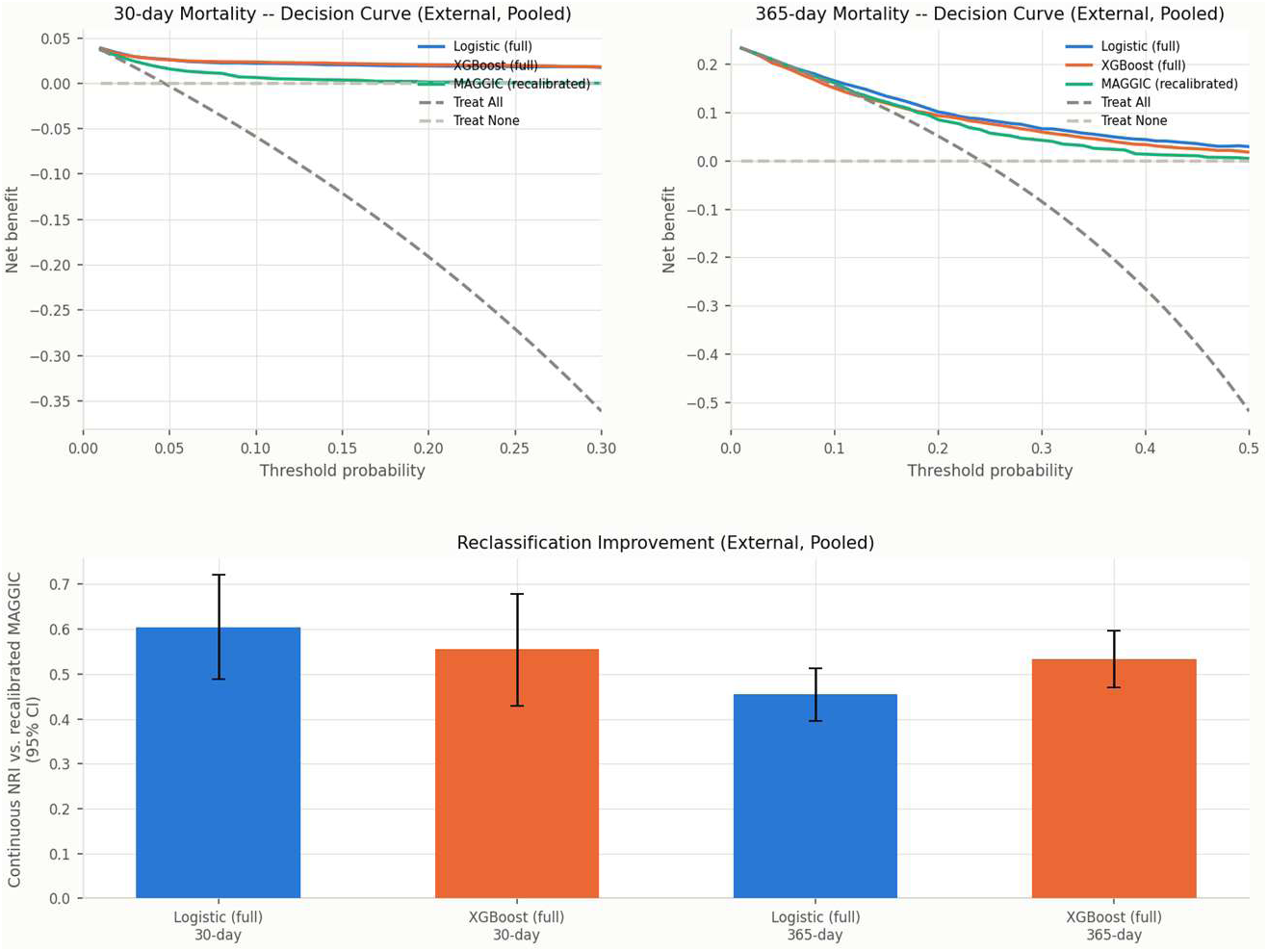
Clinical Utility: Decision Curve Analysis and Reclassification. Clinical utility. Decision-curve (net-benefit) analysis for the ML models versus recalibrated MAGGIC and the treat-all / treat-none references, with reclassification metrics, for both outcomes.

**Table 3.** Clinical utility and reclassification versus recalibrated MAGGIC. NB, net benefit at a 10% threshold probability (decision-curve analysis); NRI, net reclassification improvement; IDI, integrated discrimination improvement. 95% CIs in parentheses. All improvements are relative to recalibrated MAGGIC.

| Outcome | Model | NB @10% | Continuous<br>NRI | Categorical<br>NRI | IDI |
| --- | --- | --- | --- | --- | --- |
| 30-day | XGBoost | 0.023 | 0.556 (0.43–<br>0.68) | 0.206 (0.14–<br>0.28) | 0.224<br>(0.19–<br>0.26) |
| 30-day | Logistic | 0.022 | 0.604 (0.49–<br>0.72) | 0.220 (0.15–<br>0.29) | 0.218<br>(0.19–<br>0.25) |
| 1-year | XGBoost | 0.150 | 0.534 (0.47–<br>0.60) | 0.061 (0.03–<br>0.09) | 0.086<br>(0.07–<br>0.10) |
| 1-year | Logistic | 0.165 | 0.455 (0.39–<br>0.51) | 0.071 (0.04–<br>0.10) | 0.078<br>(0.07–<br>0.09) |

### Head-to-head against region-specific scores and robustness

Beating a Western-derived score could be dismissed as an unfair comparison; the ML models also outperformed both scores purpose-built for this population (Singapore HF Score and OPTIMIZE-HF-Asia) by a similar margin, the strongest available form of the ‘better than existing tools’ claim. The advantage was stable under every stress test. Under temporal validation (train pre-2021, test 2021-onward), XGBoost 30-day AUROC was preserved and indeed higher on contemporary admissions (0.881, 95% CI 0.817–0.938; 1-year 0.765), indicating the model does not degrade across the ARNI/SGLT2i practice shift (Supplementary Table S2). Multiple-imputation results differed from the primary median-imputation results by <0.01 AUROC (Table S4). Out-of-bag optimism-corrected internal AUROC tracked the held-out test performance closely (XGBoost 30-day: 0.832 corrected vs 0.820 actual; 1-year 0.763 vs 0.745), arguing against overfitting (Table S5).

### Confounding by indication

Discharge medications ranked among the strongest predictors, raising the concern that the model might be reverse-engineering illness severity from prescriptions. Removing all eleven medication variables reduced external 30-day AUROC from 0.862 to 0.774 (95% CI 0.743–0.804) for XGBoost and to 0.750 (0.717– 0.783) for logistic regression. Both intervals overlap recalibrated MAGGIC (0.754, 0.721–0.785), so at 30 days the medication-free models are no longer distinguishable from the best established score: the medication block carries essentially all of the short-term advantage, and the non-medication predictors alone do not outperform MAGGIC. The picture differs at one year, where the medication-free models retained a clear advantage (XGBoost 0.735, logistic 0.733, vs 0.699 for recalibrated MAGGIC). Because non-prescription of a beta-blocker or loop diuretic may encode a clinician’s judgement that a patient was too unstable to treat rather than an independent biological signal, we report both the full and medication-free models and do not interpret medication SHAP effects causally; residual confounding cannot be excluded. Both the full and medication-free models are therefore reported, and medication SHAP effects are not interpreted causally.

### Fairness across subgroups

Discrimination was broadly consistent across age, sex, ethnicity, and ischaemic aetiology, with no systematic large disparity (Figure 5; Supplementary Table S3). As expected, small subgroups produced correspondingly wide confidence intervals: the striking external 30-day AUROC in Malay patients (0.948– 0.959) rests on only 27 events, the Eurasian subgroup contributed a single 30-day death, and no subgroup discrimination estimate is reported for it. These point estimates are reported with their uncertainty rather than highlighted as evidence of superior subgroup performance.

### Interpretability

SHAP analysis showed that the model’s drivers are clinically coherent rather than artefactual. For 30-day mortality, the leading predictors were beta-blocker^17^,^18–20^ and loop-diuretic^21^ prescription, serum sodium, age, and urea; for 1-year mortality, age, urea, body-mass index, sodium, and LVEF led. All directions matched HF pathophysiology: hyponatraemia, azotaemia, reduced LVEF, anaemia, low blood pressure (a marker of pump failure in HF, not health), and older age all increased predicted risk^22^. The protective direction of diuretic prescription plausibly reflects clinical suitability for therapy as much as a direct treatment effect: patients not prescribed a diuretic despite an HF admission may be unable to tolerate diuresis because of acute kidney injury or hypotension, both markers of a poorer trajectory. Higher BMI was associated with lower predicted risk in both windows (the ‘obesity paradox’), consistent with prior analyses of this cohort. Critically, SHAP dependence plots for sodium and urea revealed sharp nonlinear threshold relationships rather than straight lines, direct visual evidence against the linear-additive assumption embedded in MAGGIC (Figure 4). Capturing those nonlinearities did not, however, translate into better discrimination on its own: the elastic-net logistic model, which cannot represent them, matched XGBoost on every external metric. The threshold effects therefore describe what the model has learned about these predictors, but the advantage over MAGGIC cannot be attributed to nonlinearity alone.

**Figure 4.**
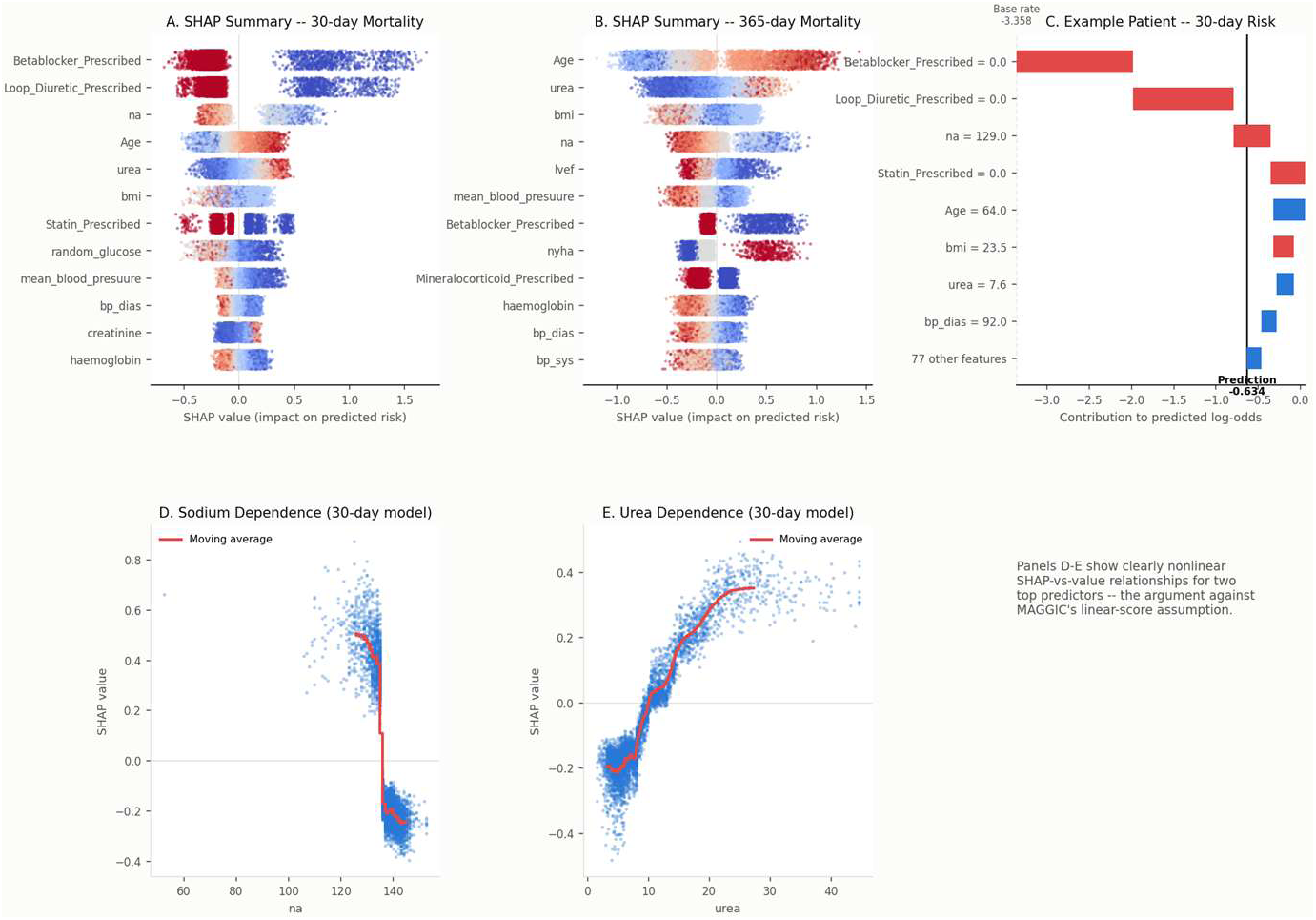
Model Interpretability (SHAP, External Validation) Interpretability. SHAP global importance rankings and dependence plots (e.g. sodium, urea) demonstrating clinically coherent, nonlinear threshold effects underlying the XGBoost predictions.

### A minimal bedside model

Sequentially adding SHAP-ranked predictors showed that a compact 10-variable model (beta-blocker and loop-diuretic prescription, sodium, age, urea, BMI, statin prescription, mean and diastolic blood pressure, and NYHA class) reproduced the full 85-feature 30-day external discrimination (AUROC 0.865 vs 0.862). One-year mortality required more variables to converge (∼30 features to reach 0.745). This supports a simple, deployable bedside instrument for short-term risk without the full data panel (Supplementary Table S7). At the point of care, a missing input would default to the same logic used throughout model development: continuous predictors to the training-set median and categorical predictors to an explicit ‘Missing’ category; no separate deployment-time imputation was developed, so any bedside implementation must retain the stored training-set statistics alongside the model. The intended implementation is a standalone risk calculator (web- or app-based) into which a clinician manually enters the ten input variables at the point of care; it does not require electronic health record integration or automated data extraction.

### From prediction to action: correctable GDMT gaps

The clinically distinguishing result is that the model localises modifiable risk. Among externally predicted-high-risk patients with reduced ejection fraction (LVEF <40%), 99.9% were missing at least one of the four GDMT pillars for the 30-day model (99.2% for the 1-year model), with a mean of 3.08 of 4 pillars absent. Because GDMT uptake was low across the entire registry, this proportion is uninformative in isolation: 95.7% of all 4,260 external HFrEF patients were missing at least one pillar, so the binary measure is close to saturated. The informative contrast is the depth of undertreatment. High-risk patients were receiving a mean of 0.92 of the four pillars, against 2.10 in model-flagged low-risk patients (n=3,438) and 1.87 across all external HFrEF patients (difference between low- and high-risk groups 1.18 pillars, 95% CI 1.11–1.24). The separation is starkest at the extreme: 36.3% of high-risk patients were receiving none of the four pillars, compared with 2.0% of low-risk patients, an approximately 18-fold difference. The model therefore does not simply inherit a low base rate of prescribing; it concentrates the most profoundly undertreated patients (Supplementary Tables S6 and S11). The single largest gap was SGLT2i (98.7% untreated in the 30-day high-risk group), plausible given it is the newest pillar, followed by mineralocorticoid-receptor antagonists (90.0%). Because most of the accrual window predates the 2021 guideline recommendation for SGLT2i in HFrEF, we stratified the gap analysis by admission era. Among 2021-onward external admissions the SGLT2i gap remained 95.7% and the any-pillar gap 99.6%, with mean pillars absent essentially unchanged (3.09 vs 3.07 pre-2021), indicating that the gap is not an artefact of calendar time (Supplementary Table S10). In short, the model does not merely rank risk. Among the patients it flags, it identifies a therapeutic deficit that is both specific and disproportionately deep relative to the cohort it was drawn from (Supplementary Tables S6, S10 and S11).

To distinguish contraindicated non-prescription from potentially remediable gaps, we reviewed documented reasons for non-prescription across the full cohort for the three pillars with structured reason-coding (ACEi/ARB/ARNI, beta-blocker, and MRA; comparable coding was not available for SGLT2i, the largest single gap identified above). Renal causes (acute-on-chronic kidney disease, chronic kidney disease, renal failure, worsening renal function, acute kidney injury, or renal impairment) accounted for 70.4% of ACEi/ARB/ARNI non-prescription (n=3,065) and 63.4% of MRA non-prescription (n=5,401); borderline blood pressure or hypotension accounted for a further 24.6% and 24.5% respectively. Beta-blocker non-prescription (n=1,096) was dominated by haemodynamic and rhythm concerns (borderline blood pressure 22.7%, hypotension 20.3%, bradycardia or heart block 19.7%). Reasons without a documented clinical contraindication were uncommon: ‘not prescribed by physician’ accounted for 2.7% of ACEi/ARB/ARNI, 7.3% of beta-blocker, and 8.9% of MRA non-users, and hyperkalaemia, often assumed to be the principal barrier to MRA use, accounted for only 1.6% of MRA non-prescription (Supplementary Table S8). This chart review suggests that most GDMT gaps in this cohort reflect appropriate clinical caution around renal function and haemodynamics rather than prescribing oversight; the guideline-addressable target above should be read as an upper bound on correctable gaps rather than an estimate of gaps due to clinician inaction alone.

### Risk stratification

Stratifying patients into model-defined risk tertiles produced clean separation of survival curves for both outcomes, with clear log-rank separation (p < 0.001); because the tertiles are defined by the model’s own predictions, this separation follows from the discrimination already reported and is shown for clinical interpretability rather than as independent evidence. Patients assigned to the high-risk tertile died substantially and visibly faster, confirming that the model’s continuous score translates into actionable, clinically interpretable risk groups rather than abstract discrimination alone (Figure 5).

**Figure 5.**
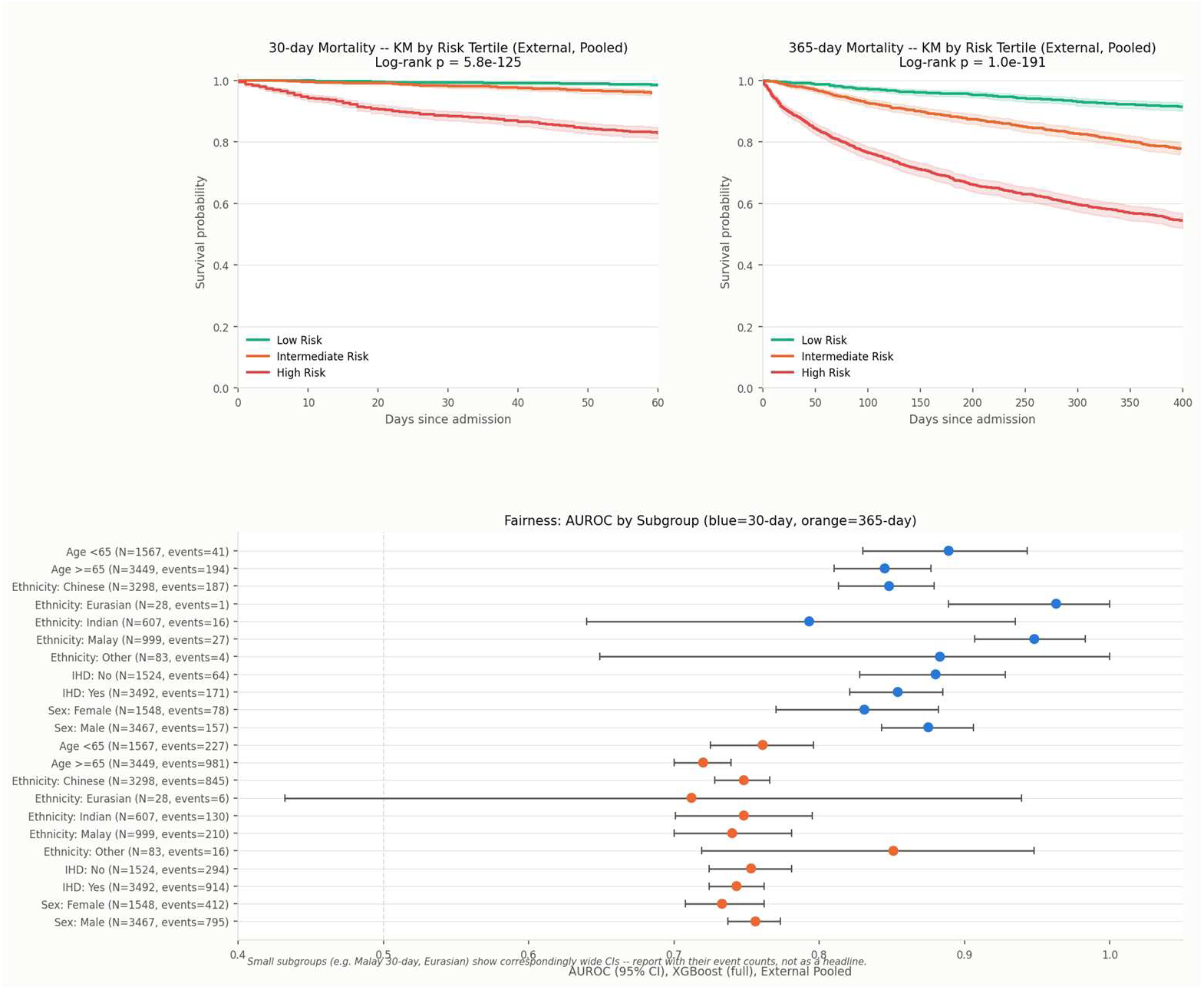
Risk Stratification and Fairness. Risk stratification and fairness. Kaplan–Meier survival by model-defined risk tertile (both outcomes) and subgroup performance across age, sex, ethnicity, and ischaemic aetiology with event counts.

## Discussion

In a large, multi-ethnic Asian HF population validated across five hospitals fully independent of model development, an interpretable machine-learning model predicted both 30-day and 1-year mortality substantially better than three established risk scores, remained well calibrated, delivered positive net clinical benefit and, most distinctively, identified a correctable guideline-directed therapy gap in nearly every high-risk patient with reduced ejection fraction. The gradient-boosted model and a transparent logistic-regression counterpart performed near-identically, and a 10-variable subset reproduced full-model short-term discrimination, indicating that the performance gain over legacy scores does not depend on either model opacity or an unwieldy data panel. Because the two model families were statistically indistinguishable on external validation, we regard the penalised logistic model as the more readily deployable of the two and report XGBoost as primary only because it supports patient-level SHAP attribution; the choice of a flexible learner is not what generates the advantage over MAGGIC.

Three features distinguish this work from prior ML HF-mortality studies. First, the comparison is fair: rather than benchmarking against a poorly scaled raw score, we recalibrated MAGGIC, the Singapore HF Score, and OPTIMIZE-HF-Asia on their linear predictors, so the ∼0.11 (30-day) AUROC advantage cannot be attributed to a miscalibrated straw-man comparator. Second, the advantage is durable: it survived temporal validation across the contemporary GDMT era (improving, not degrading, on recent admissions), multiple imputation, out-of-bag optimism correction, and removal of all medication predictors. Third, and most important clinically, the model is actionable. A more accurate risk number that changes no decision has limited value; here the same model that ranks risk also shows that 99.9% of predicted-high-risk HFrEF patients lack at least one GDMT pillar, with SGLT2i the dominant gap: converting prognosis into a specific, auditable intervention target. A chart review of documented non-prescription reasons indicates that most of these gaps reflect renal or haemodynamic contraindications rather than oversight, so this actionable fraction should be read as an upper bound; even so, a consistent minority of gaps across all three reason-coded pillars had no documented contraindication, preserving a genuine target for intervention.

The interpretability analysis characterises what the models key on, although it does not by itself account for the advantage over linear scores. SHAP dependence plots for sodium and urea show steep threshold effects rather than the straight-line relationships that MAGGIC and similar scores must assume. The model’s most influential variables (natraemia, azotaemia, age, ejection fraction, blood pressure, anaemia) are exactly the markers experienced clinicians weigh, but the model combines them nonlinearly and interactively without pre-specification.

Beyond prognostication, this kind of risk stratification offers a scalable route into clinical workflows: model-derived probabilities could flag high-risk patients at discharge for early follow-up, community heart-failure programmes, or telemonitoring, while supporting de-escalation of resource-intensive review for patients predicted at low risk3. Embedding such flags in discharge dashboards would let the actionable GDMT-gap findings above reach clinicians at the point of care rather than remaining a retrospective research observation.

### Limitations

Several limitations should be noted. First, confounding by indication is possible: discharge medications are strong predictors, and sicker patients may receive or tolerate fewer GDMT agents, so some of the apparent ‘protective’ medication signal is likely indication bias; we address this with a pre-specified medication-free model (external 30-day AUROC 0.75, confirming substantial non-medication signal) and do not interpret medication SHAP effects causally, but residual confounding cannot be fully excluded.

Second, small sites and subgroups limit precision: one external hospital contributed only 67 patients (6 thirty-day events), and certain ethnic subgroups (e.g. Eurasian, n≈28) and event counts (Malay 30-day events n=27) yield wide confidence intervals; these estimates are reported with their uncertainty and should not be over-interpreted. Third, validation, although geographically external across five hospitals, remains within Singapore, so cross-national transportability is untested. Fourth, comparator proxies may modestly disadvantage the benchmark scores: the Singapore HF Score and OPTIMIZE-HF-Asia require variables our registry captures only as proxies (e.g. arrhythmia and cerebrovascular disease standing in for atrial fibrillation and prior stroke). Fifth, the registry is retrospective and the outcome is all-cause mortality; cause-specific mortality and HF-hospitalisation endpoints were not modelled, and prospective and interventional evaluation, testing whether acting on the flagged GDMT gaps improves outcomes, is the necessary next step. Sixth, the 30-day development set contained 142 deaths against 85 encoded features, an events-per-variable ratio well below conventional recommendations; out-of-bag optimism correction indicated minimal overfitting (Table S5), but the 30-day model should be regarded as the more fragile of the two. Seventh, the cohort is predominantly HFrEF (71.9% with LVEF <40%; mean LVEF 27.6%), so performance in HFpEF is untested, and the comparison with MAGGIC, which has been separately validated in HFpEF, should not be extrapolated to that population. Finally, our registry does not capture standardised frailty assessments; frailty is closely and independently linked to HF prognosis and hospitalisation risk^23^,^24^, and residual confounding by unmeasured frailty status cannot be excluded.

## Conclusion

Developed across three hospitals and validated in five fully independent external hospitals, an interpretable machine-learning model outperformed established heart-failure risk scores for both 30-day and 1-year mortality, was well calibrated and clinically useful, and uniquely localised correctable guideline-directed therapy gaps in nearly all predicted-high-risk patients with reduced ejection fraction. Reproducible with as few as ten bedside variables, the model offers a practical prediction-to-action tool that warrants prospective, and ultimately interventional, validation.

## Data Availability

Data will be available upon reasonable request and institutional approval

## Data Availability

The analysis code, the final model coefficients, and all preprocessing constants required to reproduce predictions are publicly available at https://github.com/FigLing/exact-hf-calculator, together with a browser-based implementation of the ten-variable model that performs all computation client-side and transmits no data. Individual patient records cannot be shared publicly because the registry operates under institutional and national data-governance restrictions; de-identified data are available from the corresponding author upon reasonable request, subject to institutional and registry ethics approval.

## Sources of Funding

This study received no specific funding.

## Disclosures

The authors declare no conflicts of interest and no relevant disclosures.

## Ethics

The National Healthcare Group Domain Specific Review Board, Domain C (NHG DSRB-Domain C) determined that formal ethics review was not required (ECOS Ref: 2025-1537), given the retrospective use of fully de-identified registry data, in accordance with Singapore’s Human Biomedical Research Act 2015. Reporting follows TRIPOD+AI.

## Supplementary materials (summary)

| Item | Content | Key result |
| --- | --- | --- |
| Table S1 | Variable-level missingness | N/A |
| Table S2 | Temporal validation (train <2021 / test ≥2021) | XGB 30-d AUROC<br>0.881 |
| Table S3 | Fairness by age/sex/ethnicity/IHD | No systematic<br>disparity |
| Table S4 | Multiple-imputation sensitivity (MICE, m=5) | ΔAUROC <0.01 |
| Table S5 | Optimism-corrected bootstrap (out-of-bag) | 0.832 vs 0.820 actual |
| Table S6 | GDMT treatment-gap quantification | 99.9% ≥1 pillar missing |
| Table S7 | Minimal-feature (SHAP-ranked) model | 10 features → AUROC 0.865 |
| Table S8 | Reasons for GDMT non-prescription (chart-reviewed) | Renal causes dominate ACEi/ARB/MRA gaps; hyperkalaemia rare (1.6%) |
| Table S9 | Per-hospital discrimination and calibration (five external sites) | 30-d AUROC range 0.855–0.901; smallest site n=67 |
| Table S10 | GDMT gap stratified by admission era (pre-2021 vs 2021-onward) | SGLT2i gap 95.7% and any-pillar gap 99.6% persist post-2021 |
| Table S11 | GDMT gap base-rate comparison (high-risk vs low-risk vs all HFrEF) | 0.92 vs 2.10 vs 1.87 pillars present; 36.3% vs 2.0% on none |

## Notes

### Competing Interest Statement

The authors have declared no competing interest.

### Author Declarations

The National Healthcare Group Domain Specific Review Board, Domain C (NHG DSRB-Domain C) determined that formal ethics review was not required (ECOS Ref: 2025-1537), given the retrospective use of fully de-identified registry data, in accordance with Singapore's Human Biomedical Research Act 2015.

